# Determinants of exposure to *Aedes* mosquitoes: a comprehensive geospatial analysis in peri-urban Cambodia

**DOI:** 10.1101/2022.09.12.22278870

**Authors:** Daniel M. Parker, Catalina Medina, Jennifer Bohl, Chanthap Lon, Sophana Chea, Sreyngim Lay, Dara Kong, Sreynik Nhek, Somnang Man, Johannes S. P. Doehl, Rithea Leang, Hok Kry, Huy Rekol, Fabiano Oliveira, Vladimir Minin, Jessica E. Manning

## Abstract

*Aedes* mosquitoes are some of the most important and globally expansive vectors of disease. Public health efforts are largely focused on prevention of human-vector contact. A range of entomological indices are used to measure risk of disease, though with conflicting results (i.e. larval or adult abundance does not always predict risk of disease). There is a growing interest in the development and use of biomarkers for exposure to mosquito saliva, including for *Aedes spp*, as a proxy for disease risk. In this study, we conduct a comprehensive geostatistical analysis of exposure to *Aedes* mosquito bites among a pediatric cohort in a peri-urban setting endemic to dengue, Zika, and chikungunya viruses. We use demographic, household, and environmental variables (the flooding index (NFI), land type, and proximity to a river) in a Bayesian geostatistical model to predict areas of exposure to *Aedes aegypti* bites. We found that hotspots of exposure to *Ae. aegypti* salivary gland extract (SGE) were relatively small (< 500m and sometimes < 250m) and stable across the two-year study period. Age was negatively associated with antibody responses to *Ae. aegypti* SGE. Those living in agricultural settings had lower antibody responses than those living in urban settings, whereas those living near recent surface water accumulation were more likely to have higher antibody responses. Finally, we incorporated measures of larval and adult density in our geostatistical models and found that they did not show associations with antibody responses to *Ae. aegypti* SGE after controlling for other covariates in the model. Our results indicate that targeted house- or neighborhood-focused interventions may be appropriate for vector control in this setting. Further, demographic and environmental factors more capably predicted exposure to *Ae. aegypti* mosquitoes than commonly used entomological indices. Keywords: Aedes; saliva; geostatistical; environmental; dengue fever; Zika; chikungunya

## INTRODUCTION

*Aedes* spp. mosquitoes are globally distributed arthropod vectors that transmit devastating viruses including dengue, chikungunya, Zika, and yellow fever in addition to being competent to host multiple other pathogens (1–4). *Aedes* spp. mosquitoes are day feeders and *Ae. aegypti* and *Ae. albopictus* favor container habitats for larval stages, are well-suited for urban environments, and feed in the daytime, making them difficult targets for blanket vector control campaigns (5–9).

Much of Southeast Asia is currently undergoing urbanization, including both landscape changes and rural-urban migration. In 1980, an estimated 25% of the total population lived in areas classified as urban compared to 2019 where 50% of the total population lives in urban settings (10). In this context, there is an expansion of *Aedes* habitats coupled with an increase in co-localized human populations, potentially leading to increased *Aedes*-human contact and a resultant increase in arboviral infections.

There is increasing interest in the use of antibodies against mosquito saliva as an indicator of mosquito-borne disease risk (11–13). Much of this work has focused on *Anopheles* mosquitoes with regard to the malaria epidemiological systems, but as malaria is nearing elimination in many Southeast Asian countries, the focus is turning to the explosive arboviral outbreaks occurring in the region each year (14,15).

Several viral diseases are thought to be primarily transmitted by *Aedes aegypti* with some transmission also attributed to *Ae. albopictus*, including dengue, chikununya, and Zika viruses (16,17). While dengue surveillance systems are relatively robust in Southeast Asia, they are less so in countries reliant on clinicosyndromic approaches like Cambodia, Laos, Myanmar, or more rural areas of Thailand (18). Markers of exposure to *Aedes* mosquitoes are valuable in assessing high-risk areas where vector control can be targeted.

The allure of a tool that directly measures exposure to feeding *Aedes* (through antibodies to *Aedes* spp. saliva) is multi-fold. Currently, an array of different entomological indices are used for estimating risk of acquiring *Aedes*-borne diseases and for measuring the relative effectiveness of public health campaigns that focus on vectors (19). These indices include measuring the absence/presence of suitable habitat for the larval stages, the absence/presence or abundance of larval stages, adult abundance, and human landing catches (HLC) (20–22). Many of these measures are time- and labor-intensive and HLC is not possible in some settings for ethical and legal reasons. Furthermore, studies that attempt to look for associations between these entomological indices and risk of infection often have mixed results (23–26).

Furthermore, even in settings where *Aedes*-borne diseases like dengue fever are endemic, it is difficult to design vector control studies with sufficient statistical power to assess the impact of public health interventions (11). Dengue fever cases fluctuate drastically from year-to-year (27,28), irrespective of vector control. Plus, many infected persons exhibit mild or no symptoms and are therefore difficult to detect despite remaining highly infectious (29). As a result, trials may require extremely large target populations in order to assess the relative effectiveness of a vector control intervention.

Measuring the impact on the vector is less restrictive, more operationally feasible, and economical. Measuring vector-human contact through antibody responses to *Aedes* salivary proteins is more relevant to the epidemiological process of interest (e.g. transmission from mosquito to human) since it is a direct measure of exposure to mosquito saliva. It also does not require assumptions about associations between *Aedes* abundance and contact with humans that can vary dramatically based on climate, human behavior, and seasonality (11). A direct, quantitative measure of exposure to *Aedes* saliva is therefore useful for assessing risk of acquiring *Aedes*-borne disease, for identifying geographic spaces where transmission is likely occurring, and for measuring the impact of public health interventions.

In this study we use data from a longitudinal pediatric cohort study in Cambodia to conduct a detailed geostatistical analysis of exposure to *Ae. aegypti* mosquitoes. Recently, we demonstrated that antibody responses to *Ae. aegypti* salivary gland extract (SGE) in this setting were associated with risk of dengue virus infection (30). We also documented the presence of Zika and chikungunya infections in this setting (31), and dengue is likewise endemic. Here, we use antibody responses to *Ae. aegypti* SGE to assess demographic, household, and environmental correlates of exposure to *Ae. aegypti* mosquitoes.

## DATA AND METHODS

### Cohort data

The study location is Chbar Mon town in Kampong Spea Province of Cambodia, a peri-urban area approximately 20km from the national capital Phnom Penh (Figure 1). A cohort of 775 children were recruited for a study on dengue virus (DENV) infection and exposure to *Aedes* mosquitoes in 2018 (as described in (32)). During recruitment, patient demographic and geographic data were recorded including: age, gender, household characteristics including number of domestic water containers or use of larvicide at their house. The geographic coordinates (latitude and longitude) of each participant’s house were recorded.

**Figure 1:**
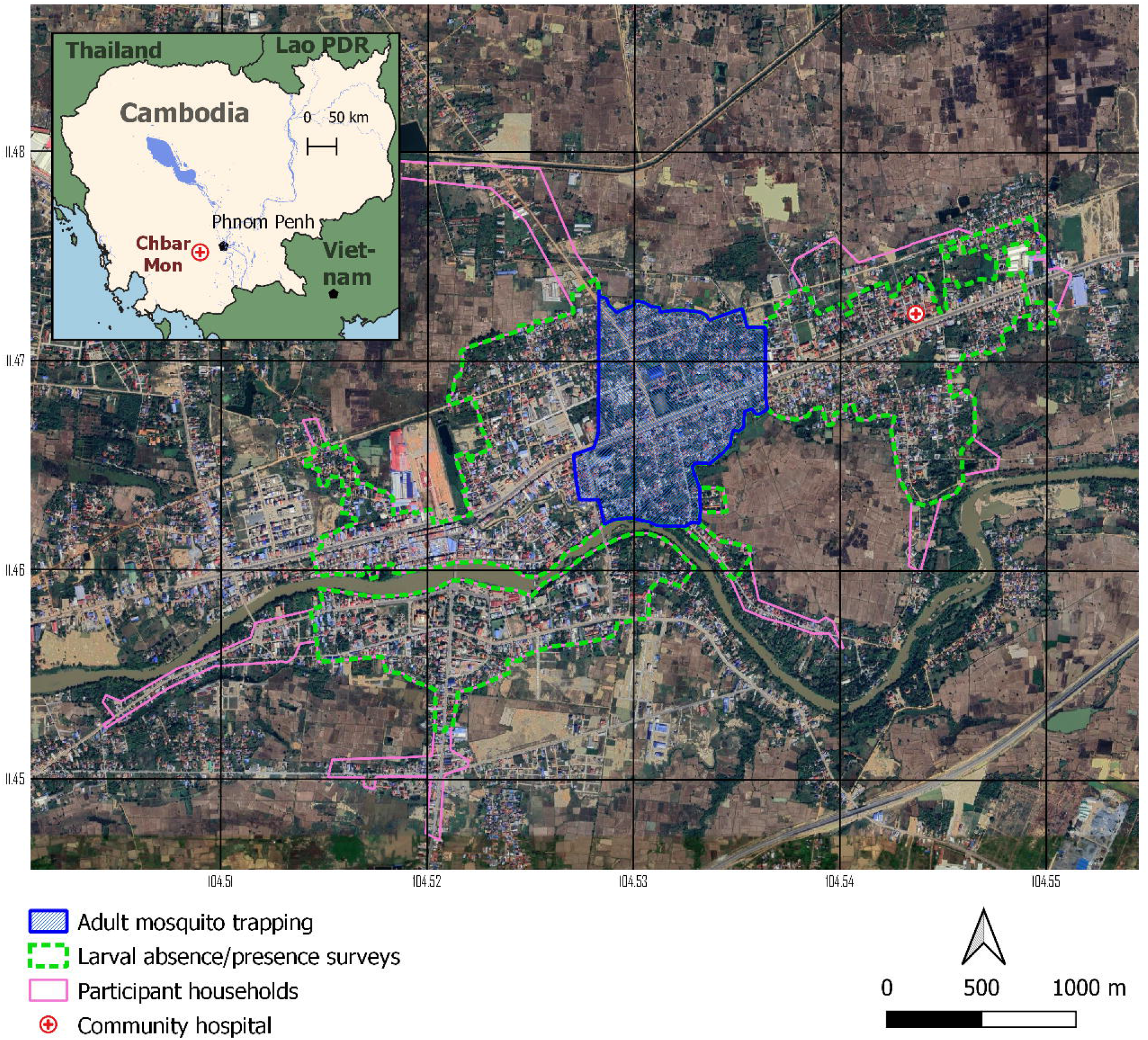
Map of study area. Inset shows location of Chbar Mon in Cambodia. Underlying map shows location of participant houses (pink diamonds); the subset of the target area where larval surveys were conducted within households (dashed green outline); and the subset of the study area where adult mosquitoes were trapped (solid blue outline).

Study staff conducted semi-annual blood sampling among the cohort: in July/August 2018; March/April 2019; July/August 2019; and March/April of 2020. Children with fever were able to present for dengue testing via rapid test and viral PCR confirmation at any time.

### House data

In a subsection of the study area (approximately 3.89km^2^), house visits were conducted twice yearly during the study period for a total of four visits. During house visits, survey staff enumerated the number of visible water containers in each house and the number of water containers infested with larvae.

### Adult abundance data

In July (rainy season) of 2018, 88 gravid traps (Biogents BG - GAT: https://us.biogents.com/bg-gat/) were set in a subsection of the target area (approximately 0.86km^2^) to trap adult *Aedes* mosquitoes to quantify adult abundance over an 8-week period of time. A 100m by 100m grid was overlaid on a central portion of the target area, encompassing a dense urban setting, a creek, and touching agricultural fields. Traps were set within each grid cell, at both indoor and outdoor settings, and were checked daily. *Ae. aegypti* and *Ae. albopictus* mosquitoes were identified and tabulated at the weekly level.

### Surface water data

A normalized flooding index (NFI) which gives an indication of surface water was extracted for this analysis (33). The data were extracted from Moderate Resolution Imaging Spectroradiometer products (MOD13Q1/MYD13Q1 250 meter AQUA/TERRA 16-day composites). The data were downloaded for each 16-day time interval (from July 2018 – May 2020) using a 250m buffer around the home of each patient in the data set. Mean NFI values in the 250m radius around each participant’s home, corresponding to the 1-month time period leading up to each patients’ hospital admission date, were used in this analysis.

### Land use/ land type data

Land cover data for study area were downloaded from Open Development Cambodia (https://opendevelopmentcambodia.net) and come from the Regional Land Cover Monitoring System at 30 by 30 m resolution. We checked the general accuracy of the land cover data using freely available satellite imagery (from Google Earth). We created 250m buffers around each participant’s house and then extracted the modal land type around each house within that 250m radius using the Zonal Histogram function in QGIS.

### Geostatistical model

A Bayesian geostatistical model was used to model individual-level exposure to *Aedes* bites regressed on the four visits’ individual level predictors, incorporating spatial correlation of the errors. The outcome variable is the log antibody response to *Ae. aegypti* salivary gland extract (SGE), hereafter referred to as exposure to “*Aedes* bites”. Linear predictors in the model included: gender, age, minimum distance from house to river, and the modal land type for an individual’s home. Linear time-varying predictors included: number of toilets and number of domestic water containers in home, use of insecticide and larvicide, home NFI, and an indicator for wet season. The sample collection periods were spaced by approximately 6 months, to assess rainy season and dry season each year. Model covariates are listed in Table 1.

**Table 1:** Table of predictor variables and their descriptions. The main outcome variable in our regressions was exposure to *Aedes* bites, measured through individual level antibody response to Ae. aegypti salivary gland extract (SGE)

| Variable | Description |
| --- | --- |
| Gender (ref: female, male) | Gender of the child, either male or female. Female is the reference group. |
| Age | Age of the child at the baseline visit. |
| Insecticide used (ref: no, yes) | An indicator for whether or not insecticide was used in or around the child's home. This data was collected at each visit. No insecticide use is the reference group. |
| Larvicide used (ref: no, yes) | An indicator for whether or not larvicide was used in or around the child's home. This data was collected at each visit. No larvicide use is the reference group. |
| Number of toilets | The number of toilets in the child's home. This data was collected at each visit. |
| Number of water containers | Number of domestic water containers in the child's home. This data was collected at each visit. |
| Modal land type (ref: urban, rice paddy, cropland) | Land cover data for study area were downloaded from Open Development Cambodia and come from the Regional Land Cover Monitoring System at 30 m by 30 m resolution. We checked the general accuracy of the land cover data using freely-available satellite imagery (from Google Earth). We created 250m buffers around each participant's house and then extracted the modal land around each house within that 250m radius using the Zonal Histogram function in QGIS. The land types were grouped into three categories: urban, rice paddy, and cropland, with urban as the reference group. |
| Distance to river | The minimum distance from a child's home to the closest point of the nearby river. This data was centered and scaled due to its range, so it is in units of standard deviations. |
| Wet season (ref: no, yes) | An indicator for if the measurement was collected during the wet season (approximately June-September), which coincides with visits one and three. The dry season is the reference group (October - May). |
| Flooding index (NFI) | An environmental index for surface water was extracted from Moderate Resolution Imaging Spectroradiometer (MODIS) products (MOD13Q1/MYD13Q1 250 meter AQUA/TERRA 16 day composites). The normalized flooding index (NFI) gives an indication of the surface water. The NFI data is for 16 day periods. In the models we used the NFI value from two periods prior to the sample collection for each child at each visit. The 1-month time period was selected because we hypothesized that it would take at least 1-month in order for there to be an immunological response from exposure to *Aedes* mosquitoes that had been spawned from flooding (time from juvenile stages, to adult, to feeding, to immunological response). A sensitivity analysis was performed to evaluate differences in results from using a buffer size of 250 versus 500 meters, lag between visit and NFI, and inclusion of other environmental indices (the normalized difference vegetation index and the enhanced vegetation index). The NFI provided the best model fit and most congruence with our |
|  | hypothesized biological process. |
| Larval container index | The larval container index variable is the percent of surveyed water containers with mosquito larvae detected within a 250 meter radius of a child's home, around the time of each visit. |
| Ave. trapped mosquitos | The average trapped mosquitos variable is the average number of adult mosquitos trapped within a 250 meter radius of a child's home, around the time of visit 1. |

Subsequent models were fit incorporating percent of surveyed water containers found to contain mosquito larvae, referred to as the larval container index, and adult abundance in a 250-meter radius around each individual’s home, on subsets of the data. Counts of larvae containing water containers were collected at times corresponding to the four antibody collection periods. Adult *Aedes* abundance was only collected in a small subset of the study area during a period corresponding the first collection timepoint. Therefore, the model including this predictor was the baseline visit model.

All model coefficients were modeled with vague normal prior distributions centered at zero, to represent the investigation’s uncertainty about the relationship between the model predictors and mosquito exposure. Statistical analyses were performed using the R statistical software system, where geostatistical models were fit using the INLA package (https://www.r-inla.org/).

The data processing, wrangling, and analytic workflow are illustrated in Figure 2. More model details can be found in the Supplemental Materials.

**Figure 2:**
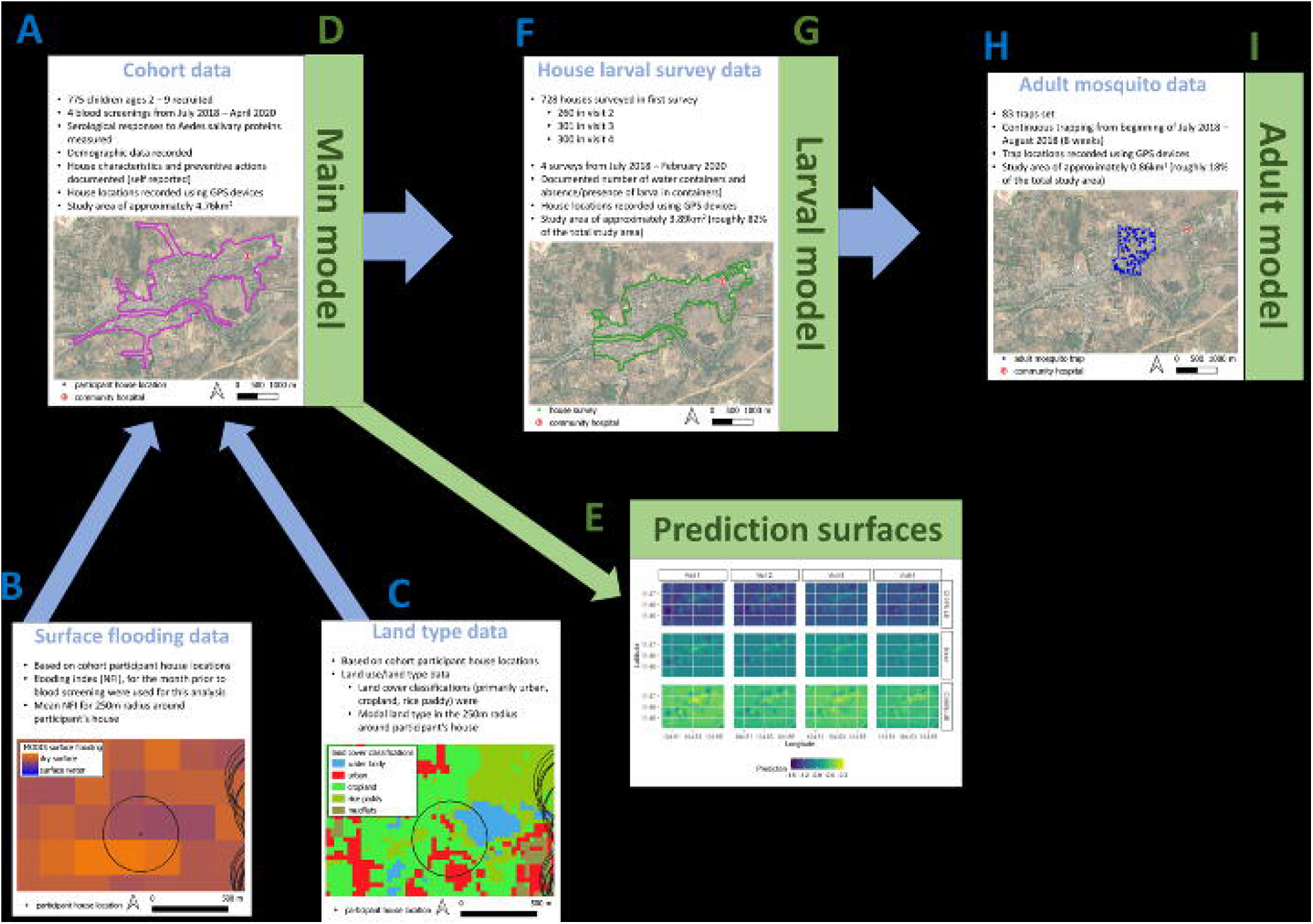
Diagram indicating data (blue letters) and analytic (green letters) steps. A.) Indicates the primary dataset used in this analysis, antibody responses among a cohort of children; B.) surface flooding data were taken from satellite imagery and merged to the cohort data based on study participant house location; C.) land type data were used to categorize participant households; D.) these data were used for the “Main model” (results in Figure 4); E.) the base model, using only the intercept, was used to generate prediction surfaces for each blood screening visit (results in Figure 3); F.) house surveys were conducted in a subset of the overall target area; G.) a geostatistical model was run, using all variables from the “Main model” but also including larvae infested containers in houses (larval model in Figure 4); H.) adult mosquitoes were trapped in a subset of the study area; I.) a geostatistical model was run, using all variables from the “Main model” but also including adult mosquito abundance (adult model in Figure 4).

### Ethics statement

The study protocol was approved by the institutional review boards at the US National Institutes of Health and the National Ethics Committee on Human Research in Cambodia. The guardians of all pediatric participants provided signed informed consent to participate in the study. The clinical protocol is available at https://clinicaltrials.gov/ct2/show/NCT03534245.

## RESULTS

### *Aedes* abundance in the study area

Both *Ae. aegypti and Ae. albopictus* were trapped in the study area. The most abundant was *Ae. aegypti* (total 436; 84.5% of all trapped *Aedes*) and this species was more abundant in indoor gravid traps throughout the 8 consecutive weeks of trapping during the rainy season (area indicated in Figure 1, details in Supplemental figure 1 and Supplemental table 1). *Ae. aegypti* mosquitoes had over 3 times the odds (3.13; 95% CI: 1.89, 5.19) of being trapped indoors when compared to *Ae. albopictus*.

### Study Population Characteristics

Approximately half of the cohort were female (50.5%) and the mean age of participants was 5.6 years. Most participant houses (60.6%) were located in urban settings, followed by unspecified cropland (35.5%) and rice paddy (3.9%). The farthest distance between any participant houses in the data is 6072m (mean 1652m) and for households not in urban settings, the farthest distance to an urban landscape was 119m (mean 33m). Approximately half of all participants reported using insecticides during the study period and less than 19% of participants reported using larvicide. The mean number of water containers per house was consistently over 3 for each visit, though slightly higher in wet seasons. As expected, mean NFI (flooding) values were highest in the wet season and lowest in the dry season (Table 2).

**Table 2:** Summary of study cohort demographics by visit. The visit columns contain the mean and standard deviation or the count and percentage for quantitative or categorical data respectively.

|  | Visit 1 | Visit 2 | Visit 3 | Visit 4 |
| --- | --- | --- | --- | --- |
| Gender |  |  |  |  |
| Female | 361 (50.49%) |  |  |  |
| Male | 354 (49.51%) |  |  |  |
| Age | 5.62 (2.23) |  |  |  |
| Land type |  |  |  |  |
| Urban | 433 (60.56%) |  |  |  |
| Rice paddy | 28 (3.92%) |  |  |  |
| Cropland | 254 (35.52%) |  |  |  |
| Distance to river (std.) | 0 (1) |  |  |  |
| Insecticide |  |  |  |  |
| No | 319 (44.62%) | 340 (53.21%) | 338 (51.52%) | 270 (43.41%) |
| Yes | 396 (55.38%) | 299 (46.79%) | 318 (48.48%) | 352 (56.59%) |
| Larvicide |  |  |  |  |
| No | 580 (81.12%) | 584 (91.39%) | 583 (88.87%) | 591 (95.02%) |
| Yes | 135 (18.88%) | 55 (8.61%) | 73 (11.13%) | 31 (4.98%) |
| Toilets | 1.36 (0.78) | 1.37 (0.95) | 1.44 (0.9) | 1.47 (1.1) |
| Water containers | 3.45 (2.38) | 3.27 (2.21) | 3.49 (2.67) | 3.29 (2.49) |
| Wet season |  |  |  |  |
| No | 0 | 639 | 0 | 622 |
| Yes | 715 | 0 | 656 | 0 |
| Flooding index (NFI) | -0.11 (0.14) | -0.27 (0.05) | -0.19 (0.09) | -0.28 (0.05) |

### Exposure ‘hotspots’ were discrete (well-separated) and persistent

Hotspots of exposure to *Aedes* bites persisted across the study duration. Figure 3 indicates the prediction surface for exposure to *Aedes* saliva for each of the time points. Clear hotspots were evident at spatial extents much smaller than 500-by-500m. Hot- and cold-spots were evident even in lower and upper credible interval maps, and across the four visits. The extent of spatial clustering (<500m and occasionally <250m) is also evident in the model variograms (Supplemental figure 2).

**Figure 3:**
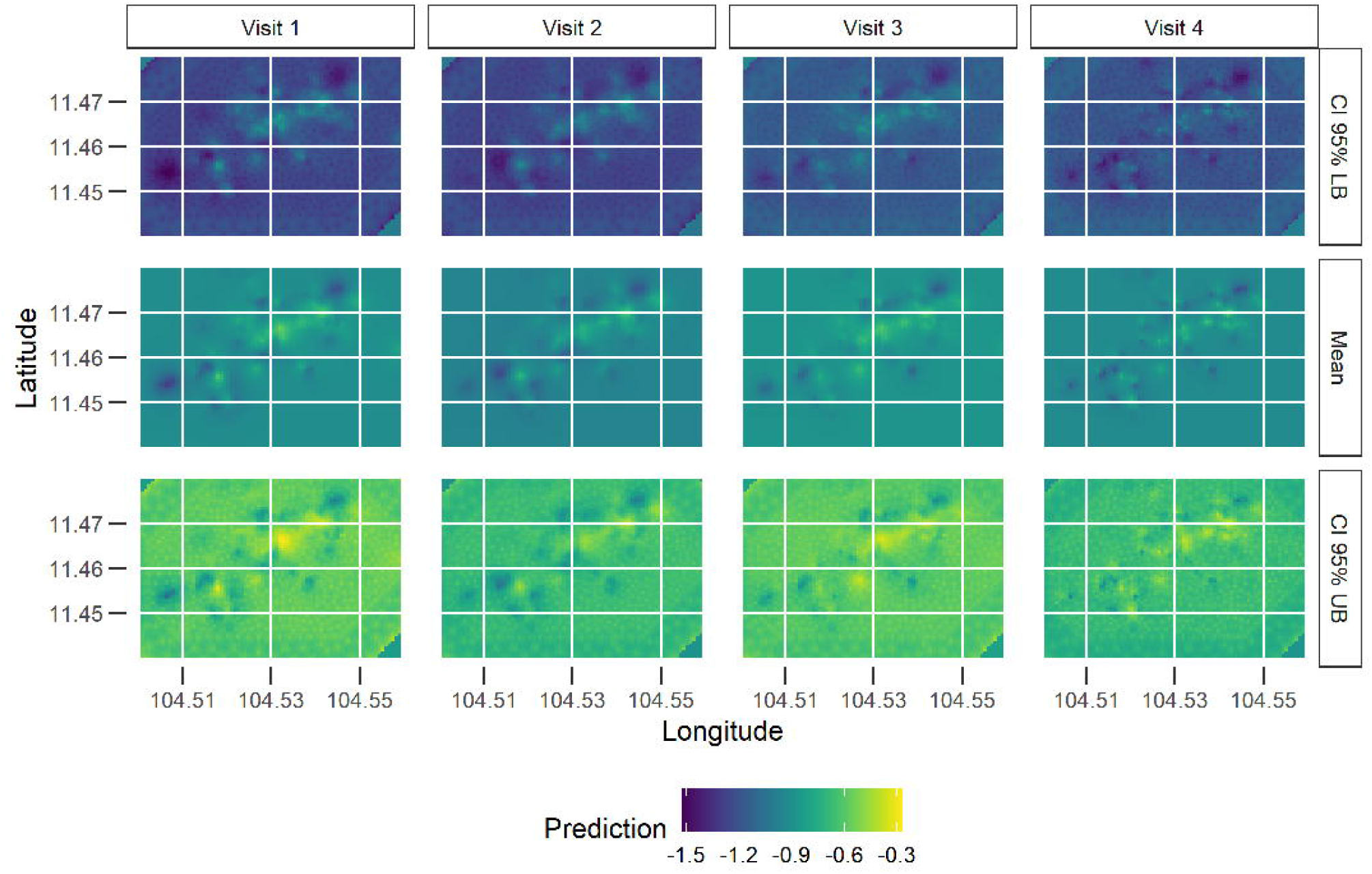
Prediction surfaces for the mean log SGE antibody response for intercept only model, by visit, with credible bounds. Row one illustrates the lower credible bounds, row two the mean predictions, and row three the upper credible bounds. Some areas consistently have higher predicted exposure (constant yellow shaded areas) relative to the rest of the study area. One area with consistently relatively high predicted exposure is located approximately within the longitudes 104.53 and 104.54 and within the latitudes 11.46 and 11.47. The lower and upper 95% Bayesian credible intervals show that these relative predicted differences in exposure are statistically significant.

### Demographic, household, and geographic predictors of exposure to *Ae. aegypti* bites

Age and gender were statistically significant predictors of high exposure to *Aedes* bites (Figure 4). The model showed that when comparing children of similar household and environmental conditions, males and older children were estimated to have lower average *Ae*. aegypti SGE antibody responses in comparison to females and younger children.

**Figure 4:**
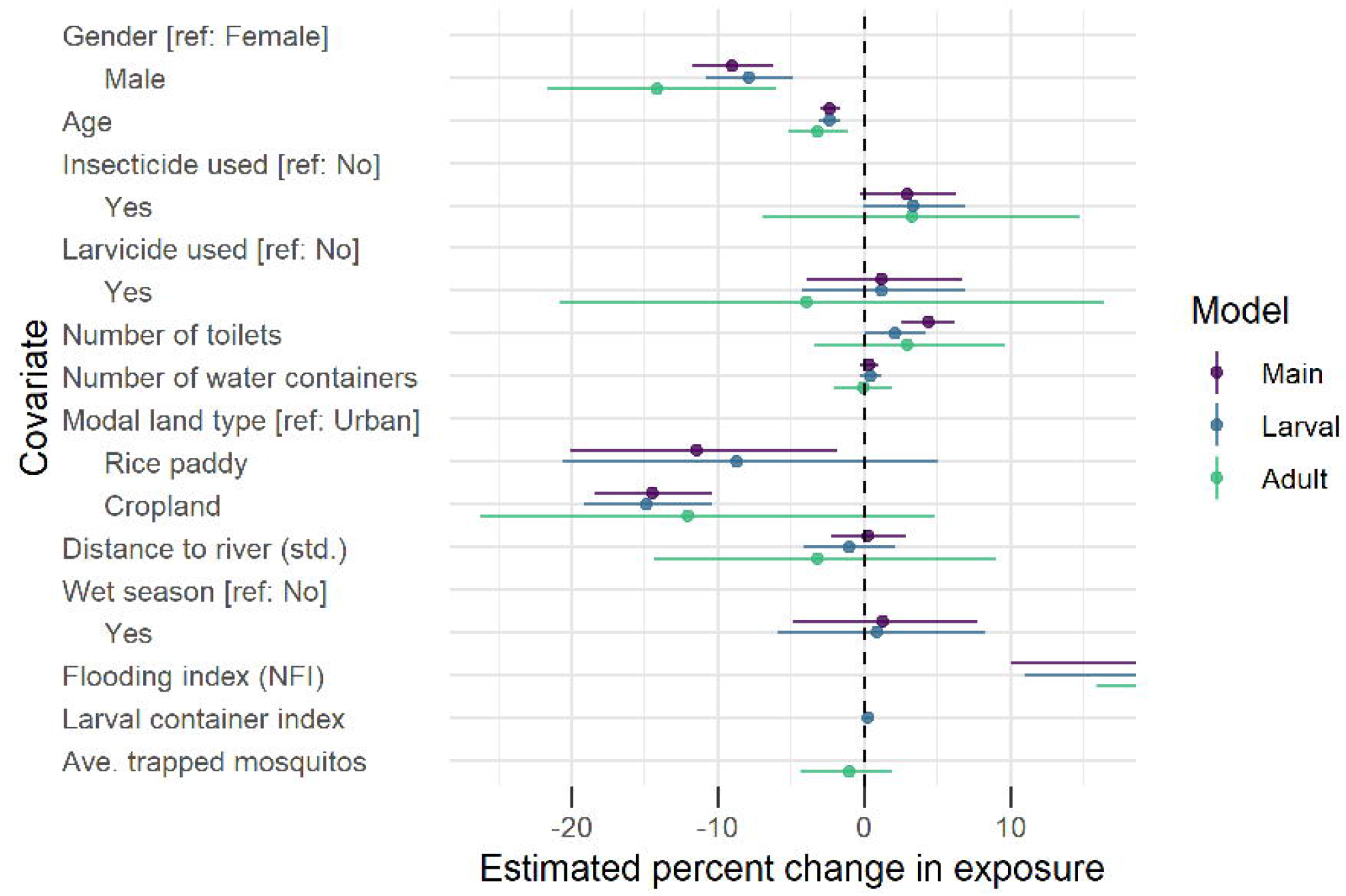
Forest plots of results from the geostatistical model predicting exposure to Aedes bites. The effect size here can be interpreted as the estimated percent change in exposure to Aedes bites. The effect size is dependent on the unit of measurement for the respective predictor variable (Table 1).

Among the household condition predictors, insecticide and larvicide use and number of water containers in a child’s house were not associated with a difference in average exposure (Figure 4 and Supplemental Table 2). When comparing children of similar demographics and environmental conditions, those with an additional toilet in their home have a mean log mosquito exposure 4.36% higher (95% credible interval of: 2.56 - 6.19). A higher mean *Aedes* mosquito exposure was detected among children who live in primarily urban areas compared to those who live in primarily cropland or rice paddy areas, controlling for flooding, season, gender, age, and household characteristics (Figure 4 and Supplemental Table 2). The model found those who lived in households with higher levels of surface flooding in the month prior to a blood screening also had higher exposure to *Aedes* mosquitoes. When accounting for flooding, land type, and the other model predictors, wet season and distance from one’s house to the river were not associated with a difference in mean exposure to *Aedes* bites.

### Larval model

The larval container index was compared to other common entomological indicators including the house index, larvae contaminated containers, and premise condition index (PCI) within a 250m radius of a study participant’s house. A plot of the correlation between these indices is in Supplemental figure 3. Mean infested containers and mean PCI had moderate correlation with the larval container index (Supplemental figure 3). The larval container index was used as a predictor in the model because it incorporates information on the number of larval infested containers while accounting for the number of surveyed water containers, which was not uniform across the study area. 98 study participants did not have any house survey data collected within a 250m radius of their house and had to be removed from the data used to build the models for this larval investigation.

Three models were compared using this subset of data: one without the larval container index; one with the larval container index; and one with the larval container index and without the environmental predictors (surface flooding (NFI), land type, distance to the river). All three models had similar estimation results and their MSE’s from cross validation showed all three models predicted approximately equally well. This is a key finding because data collection projects such as the house survey are costly and our findings suggest that the larval container index, an expensive and labor-intensive data point, can be replaced with the easily collected environmental predictors (NFI and land type) of our model.

The larval model (Figure 4) is identical to our main model but with container index included, and it had near identical estimation as the main model. Our model estimates the linear association between living in an area with a higher percentage of larval contaminated containers and exposure to *Aedes* mosquitos to be practically zero (posterior mean of 0.26 with a 95% credible interval of -0.02 to 0.54), when accounting for the other variables present in our model.

### Adult mosquito model

487 study participants did not have an adult *Aedes* mosquito trap within at 250m radius of their house and were therefore removed from the data used in the adult abundance investigation.

Again, three models were fit: one with and one without average adult abundance and one with average adult abundance but without the environmental predictors. We found these models all to have similar estimation and prediction performance. This again suggest that the costly *Aedes* abundance data can be replaced with the environmental predictors (i.e. surface flooding and land type), while also accounting for demographic and household characteristics.

The adult model in Figure 4 is the model including adult *Aedes* abundance as a predictor and it had similar estimation to the main and larval models, with increased variation due to the smaller sample size. Our findings suggest there is no linear association between nearby adult *Aedes* abundance and a child’s exposure to these mosquitos (posterior mean of -1.05 with a 95% credible interval of -4.32 to 1.87), when including the other model variables. This model also found no evidence of an association between the number of toilets in a child’s home and their exposure to *Aedes* mosquitos.

## DISCUSSION

Here we present a detailed geostatistical analysis of exposure to *Aedes* mosquitoes among a longitudinal pediatric cohort in peri-urban Cambodia. The results have practical public health relevance for *Aedes* borne-diseases, which are common throughout most of the tropical and subtropical world. Specifically, our results suggest that: 1) there are strong demographic and geographic correlates of exposure to *Aedes* mosquitoes, and 2) house-focused public health strategies are likely to be efficient at disrupting *Aedes*-human contact.

Several clear patterns emerged from our analysis. Demographic factors (age and gender) were strong and consistent predictors of exposure to *Aedes* mosquito bites. Males and older age groups had lower antibody responses to *Aedes* salivary proteins. The negative association between age and immunological response to *Aedes* salivary proteins has been described from several other studies (34–36). This be related to chronic exposure over the lifespan, with decreased antibody reaction (i.e. anergy) to *Aedes* saliva over years of exposure. At least one other study has also noted the gender effect that we describe here (34). In older age groups this effect might be attributable to gendered differences in exposure (through workplaces, etc.) but this differential exposure is not obvious for young children unless one gender is preferred to attend school (in Cambodia both genders consistently attend primary school (37)). Further work is needed to find the social or biological mechanisms behind this pattern.

Household location was likewise a consistent predictor of exposure to *Aedes* mosquito bites. Individuals who lived in agricultural areas had lower antibody responses to *Ae*. aegypti SGE than those living in urban settings. This finding is intuitive in that the primary vector of dengue virus (*Ae. aegypti*) is widely considered an urban-dwelling mosquito. Conversely, participants living in these houses are likely to visit urban settings frequently, living only a short distance from relatively dense urban dwellings (mean distance from all participant houses to urban land type was 33m). Presumably, individuals living in those households also visit schools, markets, temples and would be exposed during those visits. Yet, we are still able to detect significantly decreased anti-*Ae. aegypti* SGE antibody levels in children living in croplands and rice paddy fields when compared to those living in urban settings. Likewise, individuals who lived in households with recent surface flooding (NFI) had higher exposure to *Aedes* mosquito bites. Water bodies are important for the larval stages of mosquitoes (including *Aedes*). This finding, in combination with the findings about living in agricultural settings, strongly suggests that households are important places of exposure to *Aedes* mosquitoes. While schools are often also the targets of vector control interventions in Southeast Asia, the data are mixed as to how effective these interventions are in reducing disease (38–40). Community-based interventions should be locally tailored.

In our study we found that hot- and cold-spots of exposure to *Aedes* mosquitoes appeared stable and relatively small (<500m) throughout the study time period. The combined geostatistical results here, where small hotspots are centered on clusters of houses and are persistent across time, may be the result of short flight patterns among *Aedes* mosquitoes (often thought to not be further than 100m from place of birth (41)). Household-focused interventions such as spatial repellents may therefore be effective at disrupting human-*Aedes* exposure in this setting (and likely others).

In previous work we demonstrated that antibody response to *Aedes* salivary proteins is a significant predictor of dengue infection (30). Several other studies have likewise sought to use antibody responses to *Aedes* saliva as a predictor of dengue infection, with conflicting results (35,42–45). We hypothesize that these conflicting results are related to the relative rarity of accounting for prior DENV infection at baseline in addition to detection of clinically inapparent DENV cases. Longitudinal studies are expensive and logistically difficult, but with validated markers for exposure and disease risk, *a priori* screening of a community at the beginning of a rainy season followed by an intervention could potentially prevent *Aedes*-borne disease. This is based on the premise that disrupting human-*Aedes* exposure is likely to lead to disruption of dengue virus transmission. Zika and chikungunya viruses have also been detected in our study setting (31,46,47) and disrupting human-*Aedes* contact may likewise protect against these *Aedes*-borne diseases.

Finally, use of a biomarker of exposure to *Aedes* mosquitoes may facilitate public health intervention studies that could rely on direct measures of exposure to *Aedes* mosquitoes rather than disease outcomes, substantially lowering needed sample size numbers and cost (32). We tested models that incorporated standardized entomological indices including proximity to larvae-infested water containers or to traps with greater abundance of adult *Aedes* mosquitoes. Neither of these indices predicted antibody responses to *Ae. aegypti* SGE. Studies that use symptomatic dengue infections as the primary outcome often require extremely large cohort sizes in order to ensure a statistically significant difference between treatment and control arms. Reported dengue cases often fluctuate markedly from year to year, even in hyperendemic settings and the same is true for other *Aedes*-borne diseases. Measuring the impact of vector-focused public health interventions by using a direct measure of exposure to *Aedes* saliva can alleviate this obstacle to much needed research on vector control for mosquito-borne diseases.

## Data Availability

All data produced in the present work are contained within the manuscript

## AUTHOR CONTRIBUTIONS

Conceptualization: DMP; CM; FO; VM; JEM

Methodology: DMP; CM; FO; JSPD; VM; JEM

Software: na

Validation: DMP; CM; FO; VM; JEM

Formal Analysis: DMP; CM; FO; JSPD; VM; JEM

Laboratory work and analyses: JB; SC; SL; SN

Investigation: DMP; CM; FO; JSPD; VM; JEM

Resources: JEM

Data Curation: JEM; CM; DMP

Writing – Original Draft Preparation: DMP; CM; VM; JEM

Writing – Review & Editing: DMP; CM; JB; CL; SC; SL; DK; SN; SM; JSPD; RL; HK; HR; FO; VM; JEM Visualization: DMP; CM; VM

Supervision: DMP; VM; JEM

Project Administration: JEM

Funding Acquisition: JEM

## ACKNOWLEDGEMENTS

We thank the study participants and their parents in Chbar Mon town of Kampong Speu Province. We also thank the clinical staff of Kampong Speu District Referral Hospital for their patient care and their support for this project.

