## Supplemental Materials for "Determinants of exposure to *Aedes* mosquitoes: a comprehensive geospatial analysis in peri-urban Cambodia"

**Supplemental Figure 1**. Abundance of adult *Aedes* mosquitoes, by species, week, and trap location (indoors versus outdoors). *Ae. aegypti* were much more abundant than *Ae. albopictus*. Furthermore, *Ae. aegypti* were more likely to be caught in-doors whereas *Ae. albopictus* were more likely to be caught outdoors.

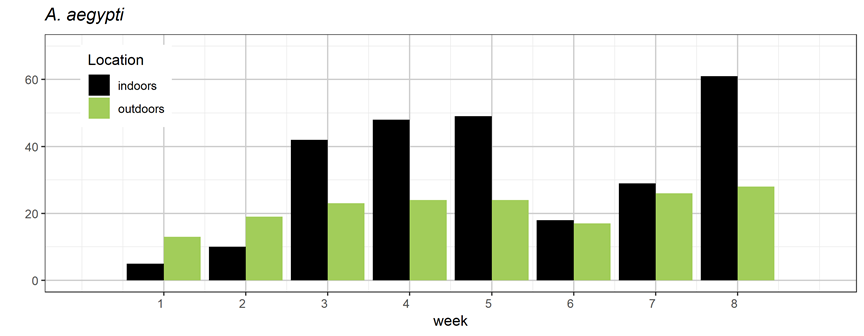

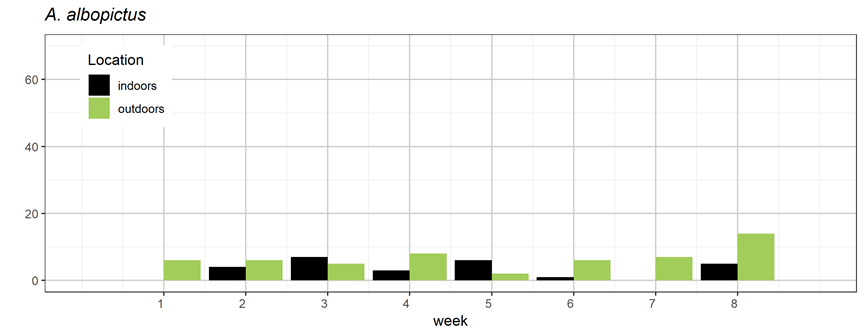

**Supplemental Table 1:** Total counts of trapped Aedes mosquitoes by species and trap location (indoors or outdoors)

|  | indoors | outdoors | total |
| --- | --- | --- | --- |
| *Ae. albopictus* | 26 | 54 | 80 |
| *Ae. aegypti* | 262 | 174 | 436 |
|  |  |  | 516 |

**Supplemental Figure 2**: Variograms from the intercept only model and full model. The variograms indicate the variance (opposite of similarity) between measurements at different spatial distances between study participant houses. Clustering is apparent at small distances, normally <500m and occasionally <250m (see Visit 1 and Visit 2 in the Intercept only models). Since there are multiple hotspots, the patterns at larger distances between participant house locations are more difficult to interpret. For example, there are clusters that are apparent in the intercept only variograms beginning at approximately 750m distance away from each other (also evident in the maps in Figure 3).

Some of the spatial clustering is removed through addition of explanatory variables in the model, apparent in the differences between the variograms in the full and intercept only models.

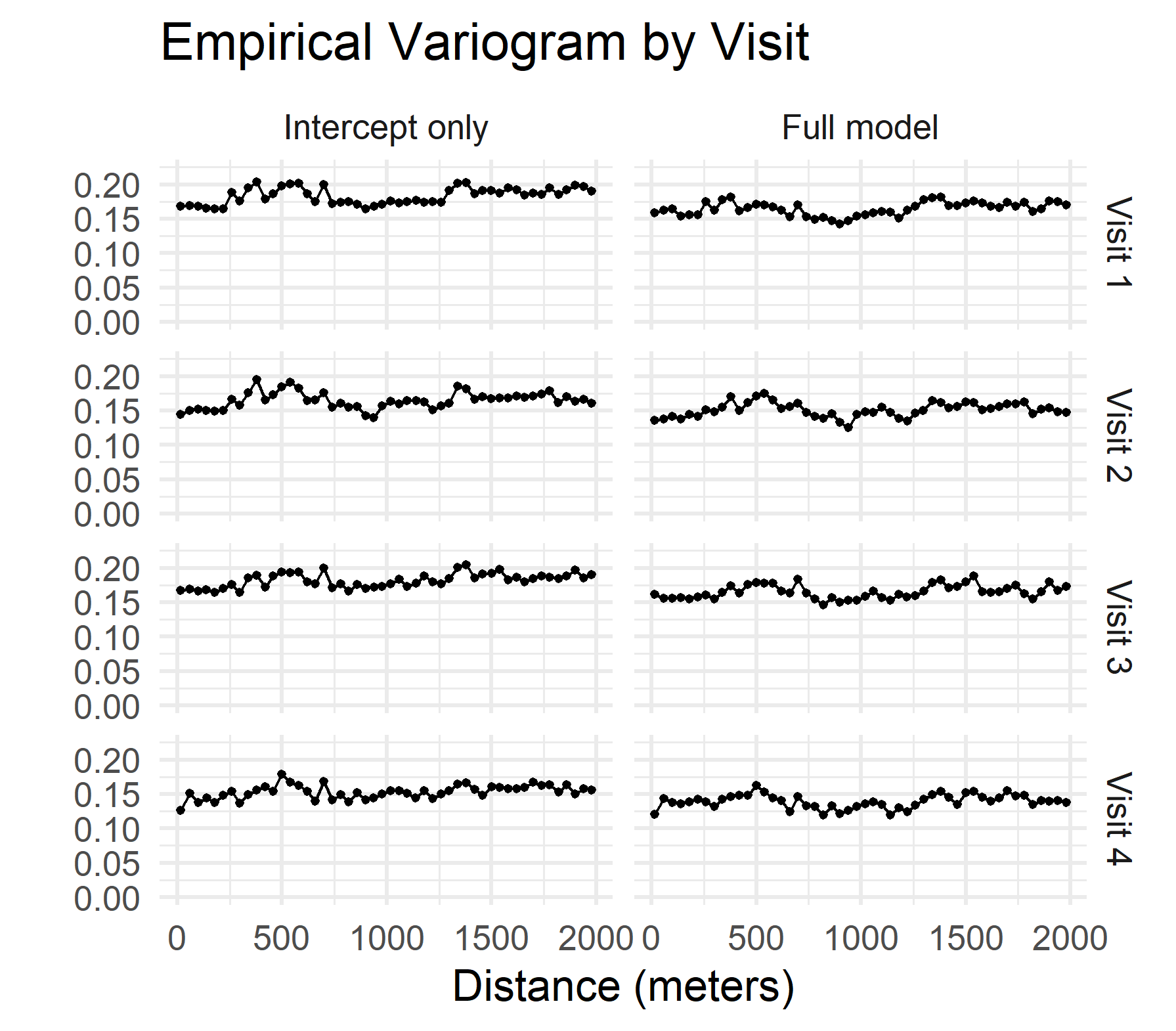

**Supplemental Figure 3**: Correlations between entomological indices (using Pearson correlation coefficients)

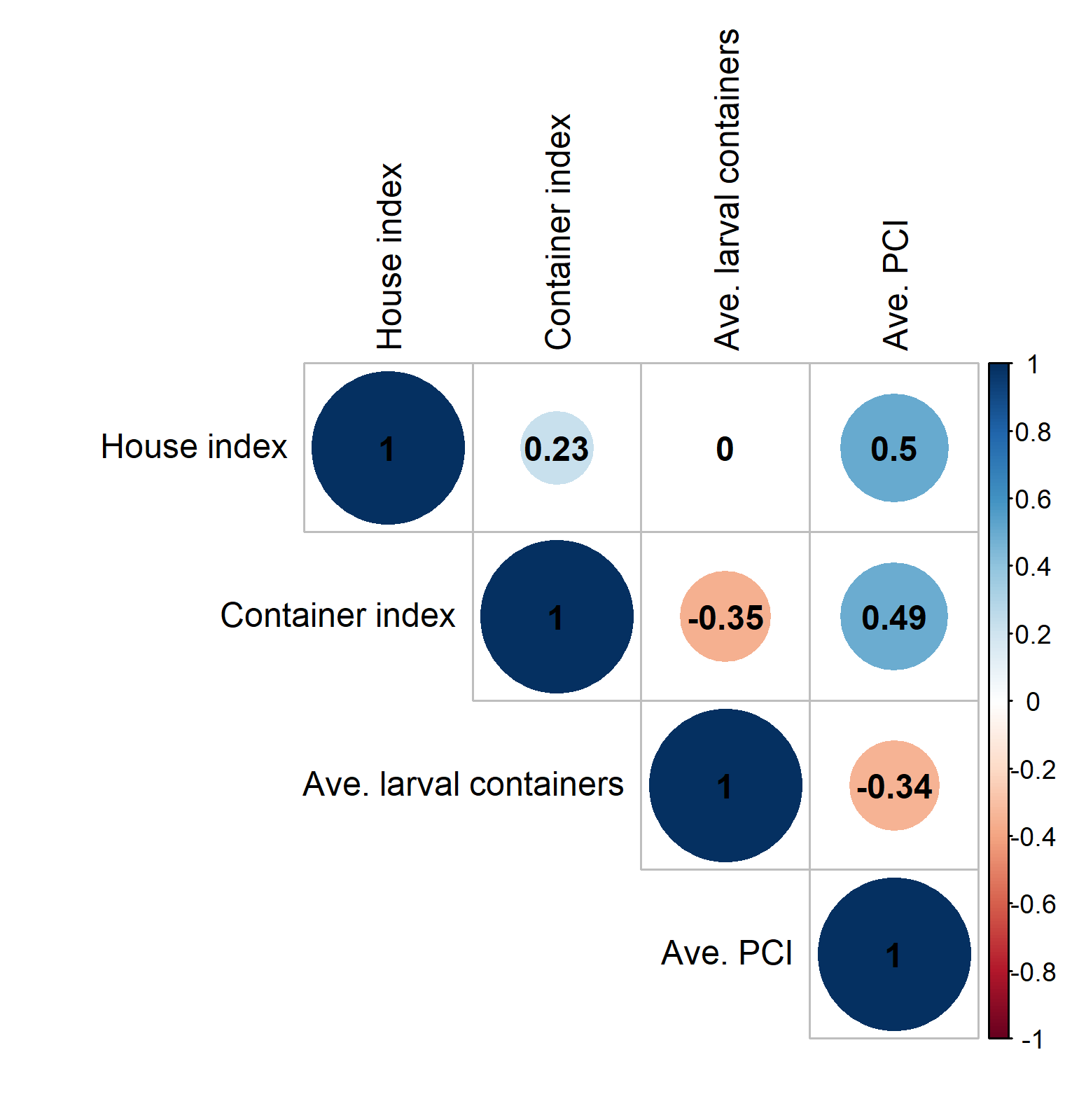

**Supplemental Figure 4**: Model results including PCI (premise condition index)

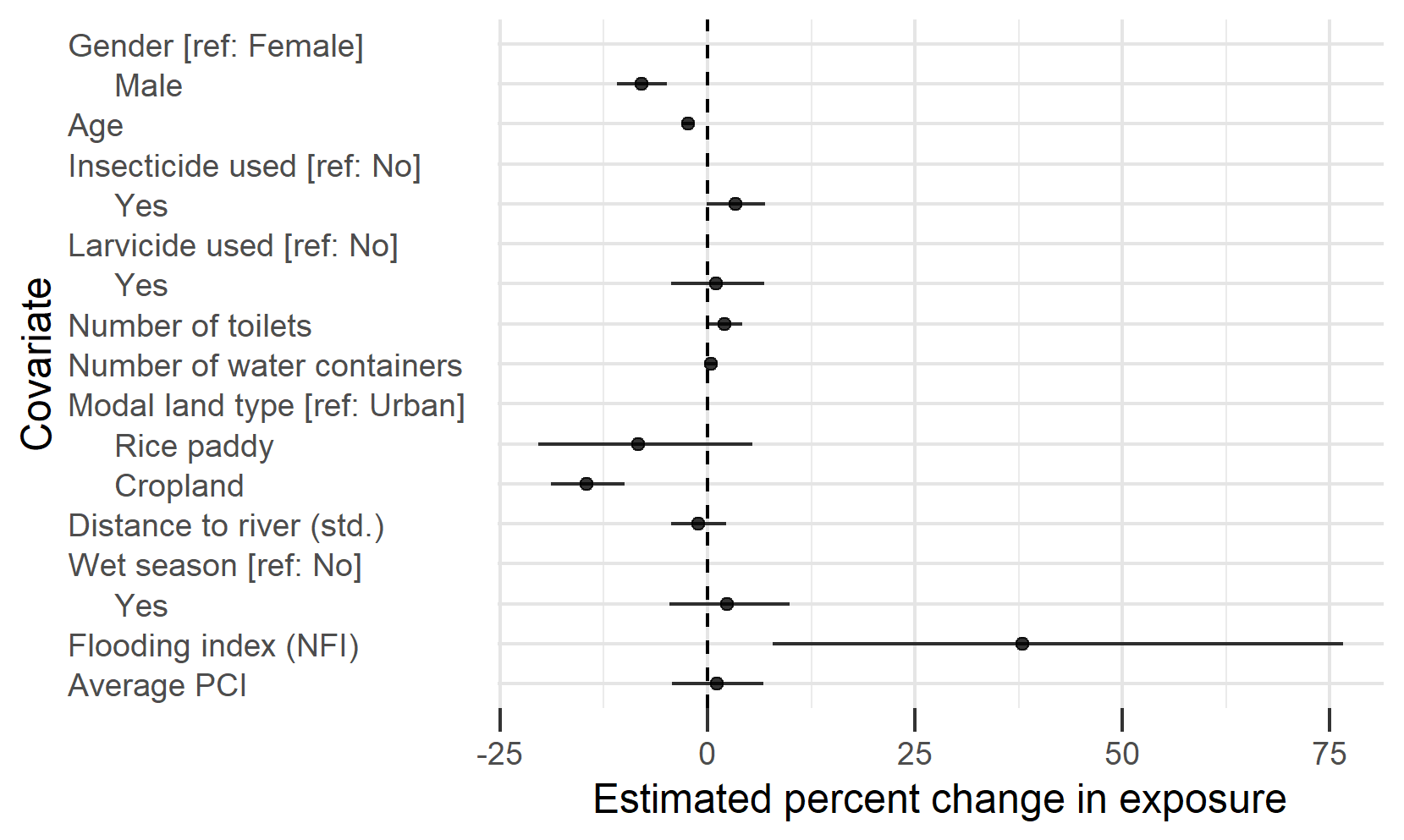

**Supplemental Figure 5**: Loess smooth of finger pick log saliva *Aedes* saliva antibody data collected on a subset of children from 4 villages in the study area. Below is the predicted exposure surface for visit 4.

From September 2020 to August 2021, an additional subset of 80 children from four randomly selected villages (20 children per village) underwent monthly finger prick collection of 200ul of capillary blood in addition to semiannual visits of venous blood collection in order to provide more granular assessments of monthly antibody kinetics of antibodies to *Ae. aegypti* SGE.

Log arbitrary ELISA units were consistent across the year with some dips in dry seasons (Supplementary figure 5). This corroborates the finding in the larger cohort that while seasonal variation in exposure exists (mean values were lower in the dry season), high responders to *Aedes* saliva exhibited high responses year-round (Figure 3).

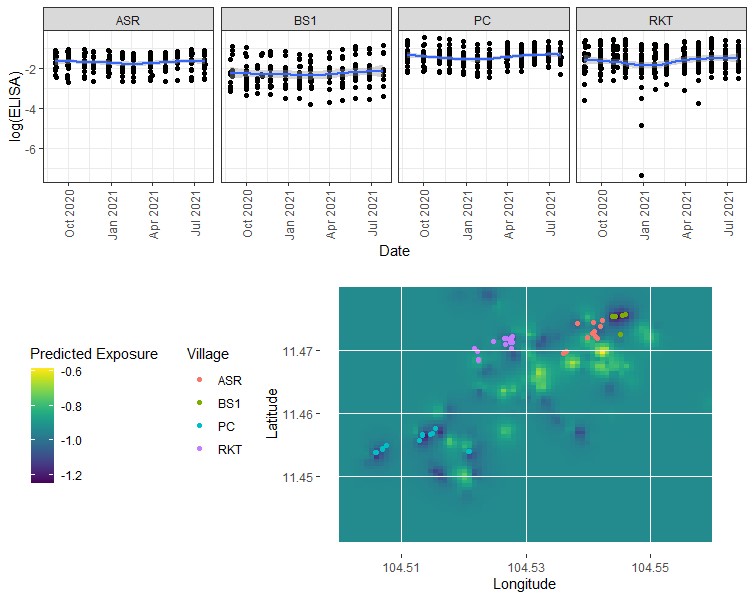

**Supplemental Figure 6**: Histogram of individual log *Aedes* saliva specific antibody levels. This distribution is useful for assessing practical significance of regression predictors. The average log antibody response is -0.91 with an interquartile range of (-1.21, -0.63).

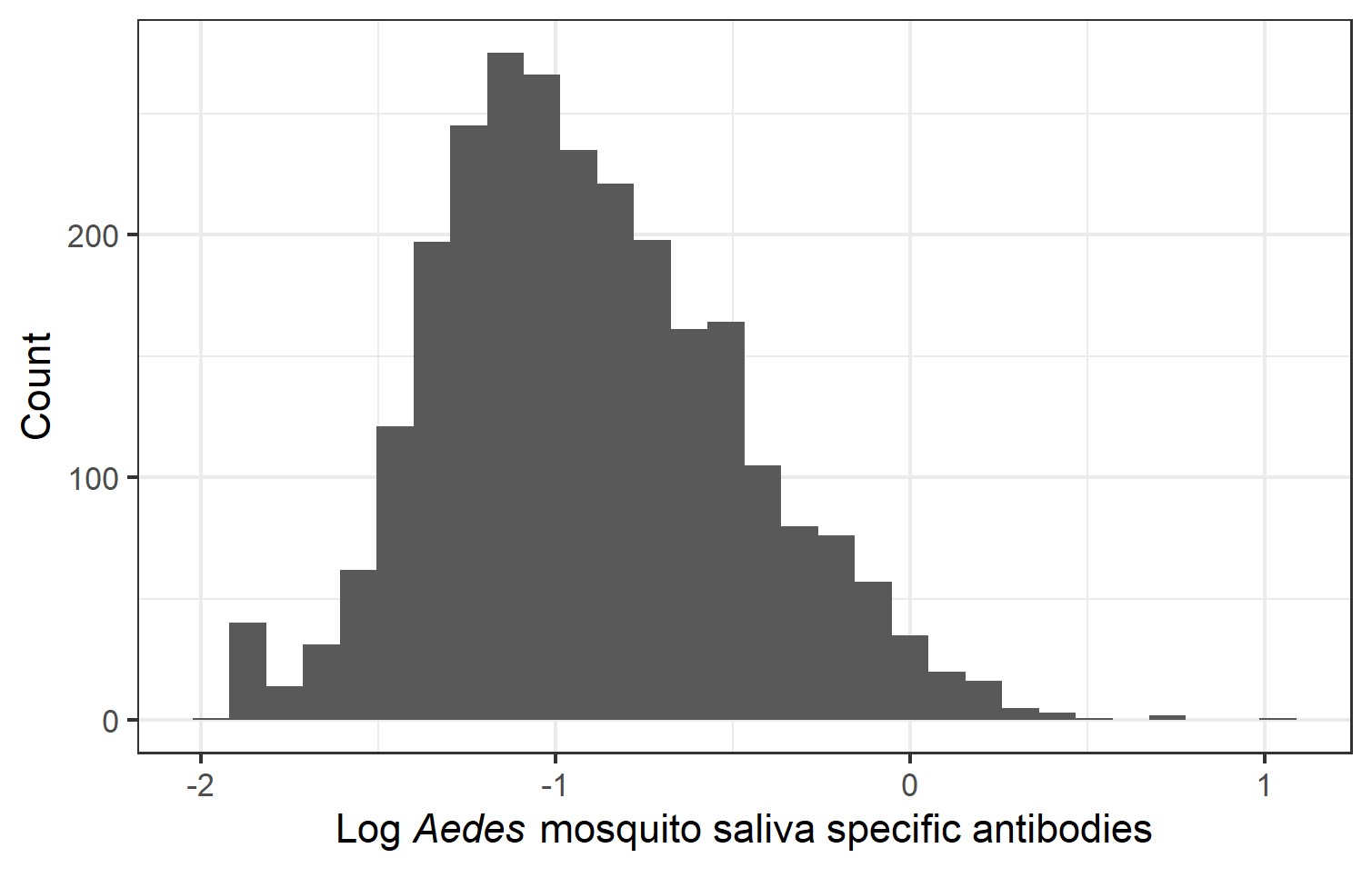

### Geostatistical model details

Some study participants had relatively high levels of *Aedes* mosquito saliva specific antibodies, so a log transformation on this response was used to reduce the range while maintaining the relative ranking of the data (**Supplemental figure 1**). A child with higher antibody levels than another will still have higher log antibody levels.

A child’s log saliva antibody levels were modeled using a spatial temporal model, specifically an Integrated Nested Laplace Approximations (ILNA) Stochastic Partial Differential Equations (SPDE) model. Spatial modeling can be very computationally intensive and the complexity increases with data size and the presence of repeat measurements. The INLA method can efficiently model geostatistical data by capitalizing on sparse precision matrices, due to deterioration of correlation between individuals due to distance, and clever utilization of Laplace approximations for normal distributions within the posterior for latent Gaussian models [1]. The SPDE approach of our INLA SPDE model indexes by spatial location instead of by study participant, and adds a continuously indexed Gaussian Random Field (GRF) term to account for spatial correlation. The mapping to space makes model computation time dependent on desired spatial resolution instead of size of data. The GRF term is modeled with normal prior centered at zero with a sparse separable precision matrix. The SPDE approach provides a connection between parameters of the precision matrix and parameters of the well understood and interpretable Matern covariance [2]. Repeat measurements can be accounted for by modeling the precision matrix as separable into the Kronecker product of a space and a time precision matrix.

There were 4 data collection time points throughout the study spaced approximately 6 months apart. This allowed for ample time for large fluctuations in an individual’s mosquito saliva specific antibodies, so there was no expectation of a correlation between observations from the same individual. As such, an individual’s repeat measurements were modeled as independent and identically distributed. Generally, our model can be written as

*y*(s_k,_t) = *X*(s_k,_ t)β + *u*(s_k,_ t) + ε(s_k,_ *t*)	(1)

where *y*(s_k,_t) is the log saliva antibody level for the individual at location s_k_ at time t for each time t; GRFs *u*(s_k,_ t) are assumed to be independent and identically distributed across times t = 1,…,4; X is the matrix of demographic, household, and environmental covariates; ε(s_k,_ t) is the Gaussian noise which is uncorrelated serially and temporally. The covariate matrix, X can be expressed as a matrix of vectors of length four, where each vector is a model covariate with all four observations for location s_k_, pertaining to individual k, specifically

*X^T^*(*s_k_,* t) = (1*,*Male(*s_k_*),Age(*s_k_*)*,*Rice paddy(*s_k_*)*,*Cropland(*s_k_*)*,*Distance to river(*s_k_*)*,*

Insecticide used(*s_k_*, t)*,*Larvicide used(*s_k_*, t)*,*Number of toilets(*s_k_*, t)*,*	(2)

Number of water containers( *s_k_*, t),Wet season(t),Flooding index(*s_k_*, t)) *.*

All coefficients in the β vector were given vague normal priors centered at zero with variance 6.25.

1. Rue H, Riebler A, Sørbye SH, Illian JB, Simpson DP, Lindgren FK. Bayesian Computing with INLA: A Review. Annual Review of Statistics and Its Application. 2017;4:395–421.

2. Bakka H, Rue H, Fuglstad G-A, Riebler A, Bolin D, Illian J, et al. Spatial modeling with R-INLA: A review. WIREs Computational Statistics. 2018;10:e1443.

**Supplemental Table 2**:

Estimated change in average individual *Aedes* mosquito saliva antibody levels, as a proxy for mean *Aedes* exposure, with 95% Bayesian Credible Interval (BCI). This table corresponds to Figure 4 in the main text. Coefficients can be interpreted as a percent change in the outcome variable.

Mean Estimate (95% BCI)

Main Model

Larval Model

Adult Model

Gender

| Female | Reference | Reference | Reference |
| --- | --- | --- | --- |
| Male | -9.00 (-11.72, -6.200) | -7.90 (-10.85, -4.85) | -14.21 (-21.69, -6.03) |
| Age | -2.34 (-3.01, -1.67) | -2.35 (-3.06, -1.63) | -3.16 (-5.16, -1.12) |

Insecticide used

| No | Reference | Reference | Reference |
| --- | --- | --- | --- |
| Yes | 2.93 (-0.32, 6.28) | 3.35 (-0.13, 6.94) | 3.28 (-7.01, 14.74) |

Larvicide used

| No | Reference | Reference | Reference |
| --- | --- | --- | --- |
| Yes | 1.21 (-3.99, 6.68) | 1.19 (-4.27, 6.96) | -3.97 (-20.86, 16.43) |
| Number of toilets | 4.36 (2.56, 6.19) | 2.09 (-0.01, 4.22) | 2.9 (-3.41, 9.58) |
| Number of water containers | 0.33 (-0.33, 0.99) | 0.42 (-0.31, 1.14) | -0.07 (-2.02, 1.92) |

Land type

| Urban | Reference | Reference | Reference |
| --- | --- | --- | --- |
| Rice paddy | -11.46 (-20.15, -1.80) | -8.70 (-20.64, 5.01) |  |
| Cropland | -14.49 (-18.4, -10.38) | -14.92 (-19.18, -10.42) | -12.04 (-26.23, 4.78) |
| Distance to river (std. dev.) | 0.26 (-2.23, 2.81) | -1.03 (-4.11, 2.13) | -3.21 (-14.40, 8.94) |

Wet season

| No | Reference | Reference | Reference |
| --- | --- | --- | --- |
| Yes | 1.25 (-4.86, 7.76) | 0.86 (-5.89, 8.24) |  |
| Flooding index (NFI) | 37.74 (10.03, 72.41) | 41.61 (10.92, 80.62) | 114.59 (15.86, 289.2) |
| Larval container index |  | 0.26 (-0.02, 0.54) |  |
| Ave. trapped mosquitos |  |  | -1.05 (-4.32, 1.87) |
